# What role do community-level factors play in HIV self-testing uptake, linkage to services and HIV-related outcomes? A mixed methods study of community-led HIV self-testing programmes in rural Zimbabwe

**DOI:** 10.1101/2024.04.16.24305916

**Authors:** Mary K Tumushime, Nancy Ruhode, Melissa Neuman, Constancia Watadzaushe, Miriam Mutseta, Miriam Taegtmeyer, Cheryl C. Johnson, Karin Hatzold, Elizabeth L. Corbett, Frances M. Cowan, Euphemia L. Sibanda

**Affiliations:** Centre for Sexual Health and HIV/AIDS Research (CeSHHAR) Zimbabwe, Harare, Zimbabwe; Faculty of Public Health and Policy, London School of Hygiene & Tropical Medicine (LSHTM), London, United Kingdom; MRC International Statistics and Epidemiology Group, London School of Hygiene & Tropical Medicine (LSHTM), London, United Kingdom; Population Services International (PSI) Zimbabwe, Harare, Zimbabwe; Department of Clinical Sciences, Liverpool School of Tropical Medicine (LSTM), Liverpool, United Kingdom; Tropical Infectious Disease Unit, Royal Liverpool University Hospital, Liverpool, United Kingdom; Global HIV, Hepatitis and STI Programmes, World Health Organization, Geneva, Switzerland; Population Services International (PSI) Washington, Washington DC, USA; Malawi-Liverpool Wellcome Trust Clinical Research Programme (MLW), Blantyre, Malawi; Faculty of Infectious and Tropical Diseases, London School of Hygiene & Tropical Medicine (LSHTM), London, United Kingdom; Department of International Public Health, Liverpool School of Tropical Medicine (LSTM), Liverpool, United Kingdom

**Author notes:** **Corresponding author** Mary K. Tumushime, CeSHHAR Zimbabwe, 4 Bath Rd, Belgravia Harare, Zimbabwe.

## Abstract

Community-led interventions, where communities plan and lead implementation, are increasingly adopted in public health. We explore what factors may be associated with successful community-led distribution of HIV self-test (HIVST) kits to guide future service delivery.

Twenty rural communities were supported to implement month-long HIVST kit distribution programmes from January-September/2019. Participant observation was conducted to document distribution models. Three months post-intervention, a population-based survey measured: self-reported new HIV diagnosis; self-reported HIVST uptake; self-reported linkage to post-test services; and viral load. The survey included questions for a composite measure of ‗community cohesion‘. Communities were grouped into low/medium/high based on community cohesion scores. We used mixed effect logistic regression to assess how outcomes differed by community cohesion. In total, 27,812 kits were distributed by 348 distributors. Two kit distribution models were implemented: door-to-door distribution only or distribution at venues/events within communities. Of 5,683 participants surveyed, 1,831 (32.2%) received kits and 1,229 (67.1%) reported using it; overall HIVST uptake was 1,229/5,683 (21.6%). Self-reported new HIV diagnosis increased with community cohesion, from 32/1,770 (1.8%) in the lowest cohesion group to 40/1,871 (2.1%) in the medium group, adjusted odds ratio (aOR) 2.94 (1.41-6.12, p=0.004) and 66/2,042 (3.2%) in the highest cohesion group, aOR 7.20 (2.31-22.50, p=0.001). Other outcomes did not differ by extent of cohesion.

HIVST kit distribution in high-cohesion communities was associated with seven times higher odds of identifying people with new HIV diagnoses, suggesting more cohesive communities may better identify those most at risk of undiagnosed HIV. Communities can learn from and adopt these participatory community-led approaches to intervention planning and implementation, which may foster cohesion and benefit public health programmes.

## INTRODUCTION

HIV self-testing (HIVST) can increase coverage and frequency of testing including among groups who would not otherwise test [1]. Since the release of the World Health Organization (WHO) guidelines in 2016, many countries have introduced HIVST and are optimising HIV testing programmes to reach those in greatest need of services at the lowest possible cost. As the scale-up of HIVST continues, there is need for evidence to inform further optimisation of HIVST distribution models to ensure the reach of individuals who remain behind.

Community-based HIVST programmes have demonstrated high impact on testing and linkage outcomes [2,3,4]. In Zimbabwe, community-based self-testing led by paid distributors achieved high uptake of HIV testing (50.3%), including among first-time testers (who comprised 36.3% of self-testers), men (46.5%) and young people under 25 years (46.2%) [2]. In addition, there was a 27% increase in uptake of antiretroviral therapy (ART) during HIVST distribution campaigns [2]. Despite its success, this HIVST kit distribution model was resource-intensive, requiring significant human and financial resources [5]. Furthermore, as testing coverage increases, the efficiency of universal testing models in terms of identification of those with unknown HIV status decreases. Therefore, there is a need for alternative, sustainable distribution models which are scalable and equitable. Such models would be particularly important for countries that are close to achieving the UNAIDS testing and treatment targets [6] or where progress among sub-populations and geographic areas is not uniform.

The community-led model has been explored as a more sustainable and empowering approach to public health interventions, with potential for lower costs. Community-led interventions, in which communities plan and lead the implementation or delivery of interventions [7], have been successfully adopted and implemented in sanitation programmes [8], dengue prevention [9], and multi-disease campaigns including HIV, malaria, hypertension and diabetes screening [10]. Due to their effectiveness, global HIV targets now include community-led approaches that advocate for the involvement of communities in planning, delivery, and monitoring HIV interventions [11]. For instance, the Global AIDS strategy 2021-2026 advocates for community-led AIDS responses and calls for 30% of testing and treatment services, 80% of HIV prevention services, and 60% of societal enabler programmes to be led by local communities and/or community organisations [12].

The success of community-led interventions can be attributed to strong local leadership and support, effective community mobilisation, community ownership, and encouraging people to have a whole-of-community rather than individual focus [8,9,10]. That each community can work together to customise their own interventions further strengthens this approach [9] and promotes community cohesion. Community cohesion, defined as the extent of connectedness and solidarity among groups in society [13], is associated with improved health outcomes [13,14] and can impact the success of community-led programmes. There is theoretical evidence to suggest that a community sense of social identity and connectedness promotes individual and group health behaviours, involvement in health-related community interventions and improved health outcomes [15,16,17]. The effect of community cohesion on HIV testing uptake in community-led interventions and subsequent linkage to prevention, treatment and care services has not been investigated.

In a trial conducted in Zimbabwe and reported separately [18], we determined the effect of community-led HIVST kit distribution on linkage to post-test services (confirmatory testing following reactive self-tests, voluntary medical male circumcision (VMMC) and pre-exposure prophylaxis (PrEP)) and self-reported recent/new HIV diagnosis. In that trial, each intervention community (cluster) was allowed to design and implement its own model of HIVST kit distribution. Here we explore the effects of the different community-led HIVST kit distribution models, levels of community involvement in planning distribution programmes and community cohesion on: (i) HIVST uptake, (ii) linkage to post-test services (confirmatory testing, VMMC and PrEP) following HIVST, and (iii) HIV-related outcomes (new HIV diagnosis and undetectable viral load). We hypothesised that communities in which distribution relied solely on distributors’ efforts (i.e., only door-to-door distribution), would perform poorer on outcomes i-iii above, compared to those in which community members actively sought and accessed HIVST kits. Furthermore, we hypothesised that closely-knit or more cohesive communities would achieve better outcomes; see Figure 1 (conceptual framework).

**Figure 1:**
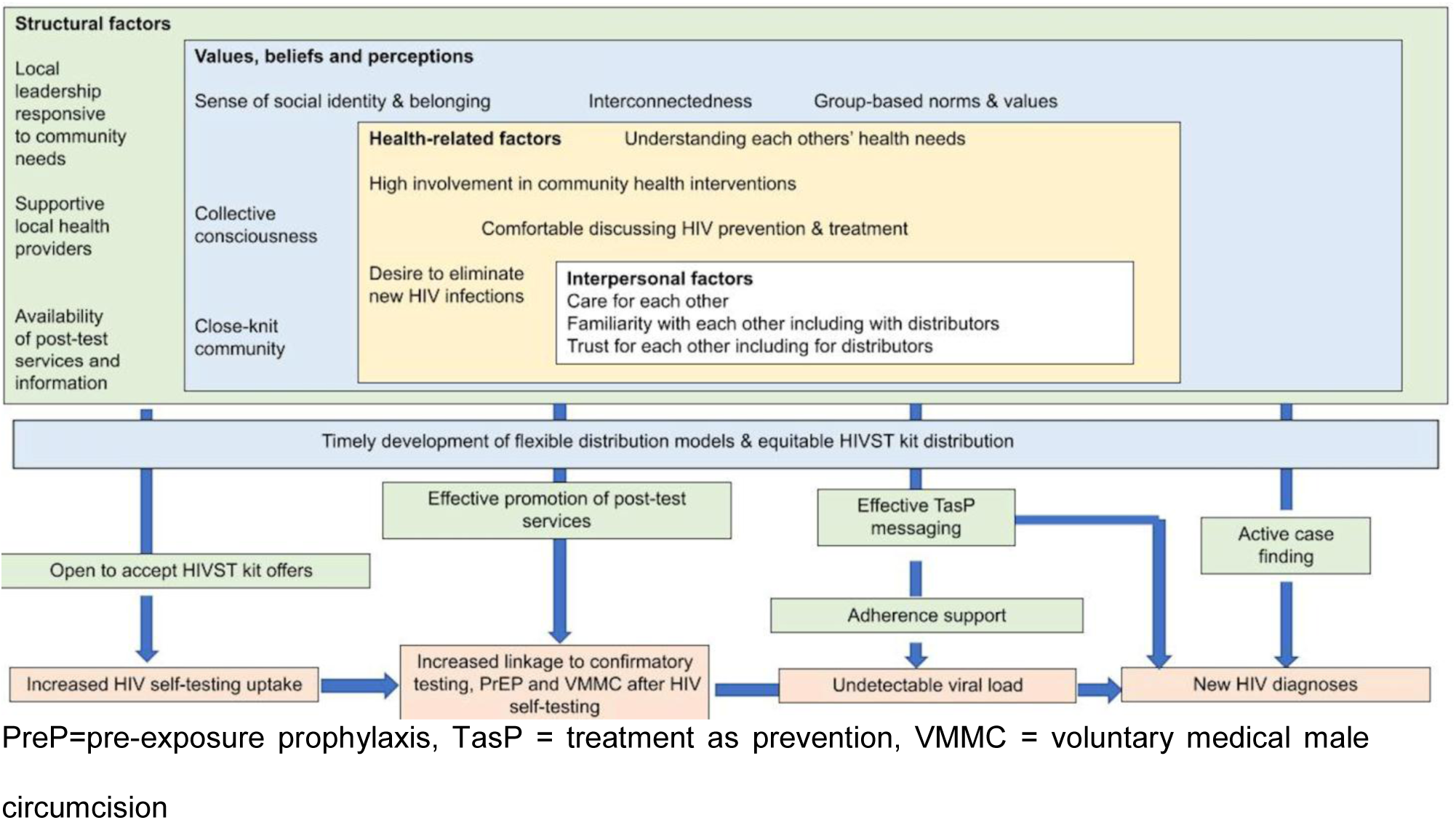
Conceptual framework – Cohesive communities achieve better HIV-related outcomes.

## METHODS

### Setting

This study was conducted as part of the Unitaid-funded HIV Self-Testing Africa (STAR) Initiative (https://unitaid.org/project/self-testing-africa-star/#en), the largest evaluation of HIVST in Africa to date, that sought to catalyse the market for HIVST and drive global scale-up. The work presented here was nested within a cluster randomised trial in rural Zimbabwe which compared HIV testing and linkage outcomes between community-led HIVST kit distribution (intervention) and community-based HIVST kit distribution led by paid distributors (comparison) [18]. Clusters/communities were defined as groups of adjacent villages headed by a local leader known as a headman (headman unit) and separated by at least 20 km. Forty headman units in 6 districts were randomised 1:1 to the study arms. This paper focuses on the intervention arm.

#### Implementation of the community-led intervention

The community-led HIVST kit distribution intervention was implemented in 20 headman units. In each headman unit, Population Services International (PSI) Zimbabwe conducted community engagement activities over a period of 2-3 weeks, firstly to introduce the concept of community-led HIVST and telling headman units that ensuring people who needed HIV treatment received it could ultimately reduce the number of new infections in their villages. Community engagement included promotion of ―U=U‖ (Undetectable=Untransmittable), where people learn that those with an HIV viral load below the limit of detection have a greatly reduced risk of onward transmission [19]. We packaged HIVST with U=U messaging. We expected that knowing comprehensive treatment can almost eliminate new HIV infections in communities and that early ART treatment can reduce HIV morbidity and mortality, would promote community members to seek HIV testing and treatment. Headman units were then invited to participate in the study and asked to design their own models of HIVST kit distribution to suit their context. Decisions about distributor selection, access to kits and/or distribution models and provision of incentives for distributors were driven by community members in the headman units and as such, distribution models were allowed to vary across the 20 communities. HIVST kit distributors identified from each headman unit were trained to promote and support HIVST and to promote uptake of confirmatory testing, VMMC and PrEP following self-testing. The local health facilities provided oversight of the distribution, supported HIVST kit storage and replenishment and provided post-test linkage services and/or referral where VMMC and PrEP were unavailable (confirmatory testing for HIV was available from all health facilities). Headman units had to adhere to regulatory requirements e.g., minimum age requirements for HIVST and non-coercive testing, which were conveyed throughout community engagement. In all headman units, distribution proceeded for 4 weeks. Headman units were given posters and flyers designed by PSI Zimbabwe to advertise HIVST availability and U=U (they decided how best these materials could be distributed/displayed for maximum effect).

Based on observations (described below), headman units implemented the following HIVST kit distribution models: (i) door-to-door distribution only or (ii) a combination of different delivery approaches including door-to-door and collection of kits directly from distributors at their homes or at various locations in the headman unit (combined HIVST distribution model). The latter model was more participatory and headman units would refine their models iteratively. Changes included community members (including distributors) forming committees to provide ongoing planning and logistical support for distribution; distributors forming pairs or groups to support each other; distributors working in villages they had not initially been assigned to, to increase coverage; and distributors taking advantage of community gatherings (e.g., meetings and sporting events) and workplaces (e.g., mines) to distribute self-test kits.

### Data collection

#### Participant observations

Participant observations were conducted by trained qualitative researchers at and between community sensitisation and planning meetings, during distributor training and during kit distribution in each headman unit to explore their progress and as part of process evaluation. Observation findings were documented using a template which captured levels of attendance and diversity of attendees (men, young people, leaders) at sensitisation meetings, levels of participation at planning meetings, how decisions were made (whether through consensus or by coercion from community leaders), degree to which headman units appeared to be cohesive and aware of community-led HIVST kit distribution and/or HIVST (ascertained through informal discussions with community members, leaders and healthcare workers), and presence in the community of promotional material (posters and flyers) about the intervention and HIVST.

#### Population-based survey

We conducted a representative population-based survey in randomly-selected households from three months after the end of HIVST kit distribution in each headman unit (08 October-30 December 2019). First, we randomly selected three enumeration areas in each headman unit, followed by random selection of one in two households. All individuals aged ≥16 years in selected households were invited to participate. The questionnaire was self-administered in the preferred language (English or two major local languages) using audio-computer assisted survey instrument (ACASI). Participants were asked about household and individual demographic characteristics, HIV testing history, experiences with HIV self-testing and linkage to post-test services. Participants were also asked to respond to a six-item measure of community cohesion validated by Lippman et al. in high HIV prevalence settings in South Africa: (i) people in this community are willing to help their neighbours, (ii) this is a close knit community, (iii) people in this community can be trusted, (iv) people in this community get along well with each other, (v) people in this community share the same values and (vi) people in this community look out for each other [20]. All items had response options of strongly agree, somewhat agree, neither agree nor disagree, somewhat disagree, and strongly disagree. Item response modelling (IRM) was used to assess and summarise the cohesion scale using a validated, one-parameter multinomial model [20]. To verify self-reports of HIV status and measure viral load, dried blood spot (DBS) samples were taken to test for HIV and viral load.

### Outcomes

For this analysis, outcomes were based on self-reports among survey respondents and viral load results. The following outcomes were compared between (i) levels of community cohesion (ii) the two distribution models headman units employed, and (iii) levels of community involvement in planning:

1. Proportion of participants self-reporting uptake of HIVST. The numerator was the number of surveyed participants self-reporting they used an HIV self-test kit to test themselves, the denominator was the total number of surveyed participants.
2. Proportion of participants self-reporting linkage to post-test services following HIVST (combined and individual measures):

i. confirmatory testing: the numerator was the number of surveyed participants self-reporting uptake of confirmatory testing following a reactive (HIV positive) self-test result, the denominator was the total number of surveyed participants who self-reported a reactive self-test result
ii. VMMC: the numerator was the number of male participants self-reporting uptake of VMMC following a non-reactive (HIV negative) self-test result, the denominator was the number of male participants who self-reported a non-reactive self-test result
iii. PrEP: the numerator was the number of participants self-reporting uptake of PrEP following a non-reactive (HIV negative) self-test result, the denominator was the number of surveyed participants who self-reported a non-reactive self-test result
3. New HIV diagnosis following HIV self-testing: New HIV diagnosis was defined as a self-reported, new provider-confirmed positive test since the start of HIVST kit distribution. The numerator was the number of surveyed participants reporting a new HIV diagnosis, the denominator was the total number of surveyed participants.
4. Undetectable viral load among people living with HIV (<1,000 copies/ml). The numerator was the number of surveyed participants with undetectable viral load, the denominator was the total number of HIV positive participants, as determined through laboratory testing of DBS samples.

### Data analysis

#### Participant observations

We described levels of community involvement in planning distribution programmes using six attributes shown in Table 1, each with three categories agreed by the research team. Information pertaining to these attributes was detailed in the template described above for each headman unit. Two researchers reviewed observation reports and independently scored headman units on each attribute, with scores ranging from 1-3, with the lowest score indicating the lowest level of community involvement. Discrepancies were discussed and resolved with consensus. Scores were totalled for each headman unit. Headman units were then categorised by terciles (low, medium, and high) indicating their level of community involvement [21]. Construction of the scale which guided categorisation was done at the beginning, and cut-off points were based on ranks, as the measure was not normally distributed. For each headman unit, distribution models were coded (1) door-to-door distribution only or (2) combined HIVST distribution model. Community involvement scores and coded distribution models were merged with survey responses.

**Table 1.**
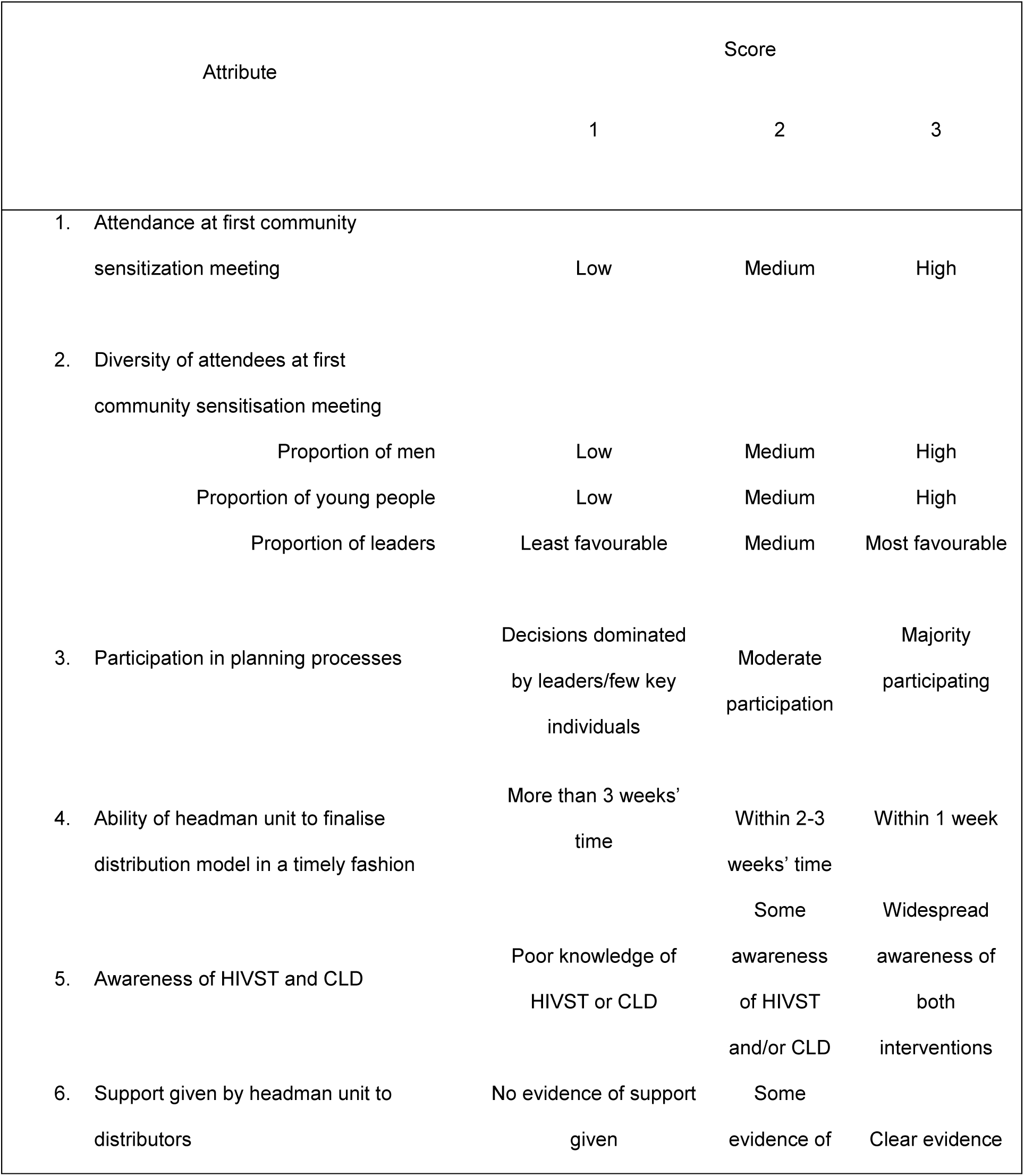
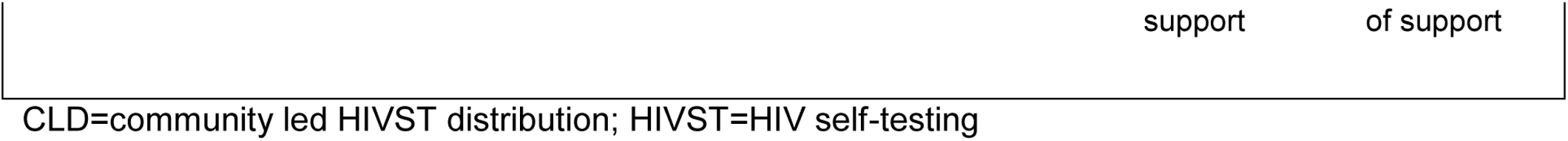
Attributes from participant observations used to determine levels of community involvement in planning distribution programs.

#### Population-based survey

Participants responded to a six-item measure of community cohesion (described above). Individual cohesion scores were calculated using the average of item responses ranging from 1 (strongly agree) to 5 (strongly disagree). Community cohesion was summarised as the median score of individuals within each headman unit, and headman units were then categorised by terciles (low, medium, and high cohesion) [21]. Construction of the scale is as above.

We used mixed effect logistic regression to assess how the outcomes above differed by distribution model, levels of community involvement and community cohesion.

Analysis used Stata v14. Before analysis, we compared similarity by distribution model, levels of community involvement and cohesion group for pre-specified variables to identify substantial differences that would need adjustment. All outcomes were analysed using mixed effect logistic regression. All models are adjusted for district, age, gender, and educational attainment. All models adjusted for the study community using a random effect. Fisher‘s exact test was used to determine if there were significant associations between the community level measures: (i) distribution model and levels of community involvement, (ii) distribution model and community cohesion, and (iii) levels of community involvement and community cohesion.

### Ethical considerations

Ethical approval to conduct this study was obtained from the Medical Research Council of Zimbabwe (ref. MRCZ/A/2323), London School of Hygiene and Tropical Medicine Research Ethics Committee (ref. 15801-1) and World Health Organization Research Ethics Review Committee (ref. ERC.0003065). The trial was registered with Pan African Clinical Trial Registry, ref PACTR201811849455568. Trained research assistants obtained written informed consent from participants prior to the survey and collection of DBS. Consenting was conducted in participants’ preferred language (English or one of two major local languages) according to standard operating procedures; this included information giving including about the study, its purpose and procedures, rights of participants, clarification of matters arising, and comprehension assessment. Informed consent forms were completed in duplicate by participants and research staff, with each retaining one copy. Where required, witnesses were present and co-signed consent forms. Participants were aged at least 16 years; a waiver of parental consent for participants aged 16-17 years was approved, given the age of consenting to HIV testing is 16 years old in Zimbabwe and survey questions were related to HIV testing.

## RESULTS

### Implementation of community-led HIVST kit distribution models and survey response rate

Implementation of the community-led HIVST kit distribution models (inclusive of community engagement, HIVST distribution and the survey) was conducted between 01 October 2018 and 30 December 2019, in 20 headman units randomised to the community-led HIVST kit distribution arm. Five headman units (5/20, 25.0%) in 3 study districts implemented door-to-door distribution only (Table 2). In the other 15 headman units (15/20, 75.0%), which were spread across all 6 study districts, a combination of different delivery approaches was used including door-to-door combined with collection of kits directly from distributors at their homes or at various locations in the headman unit (combined HIVST distribution model). Overall, 348 distributors were trained and distributed 27,812 kits, with a range of 28-159 kits distributed per distributor.

**Table 2.**
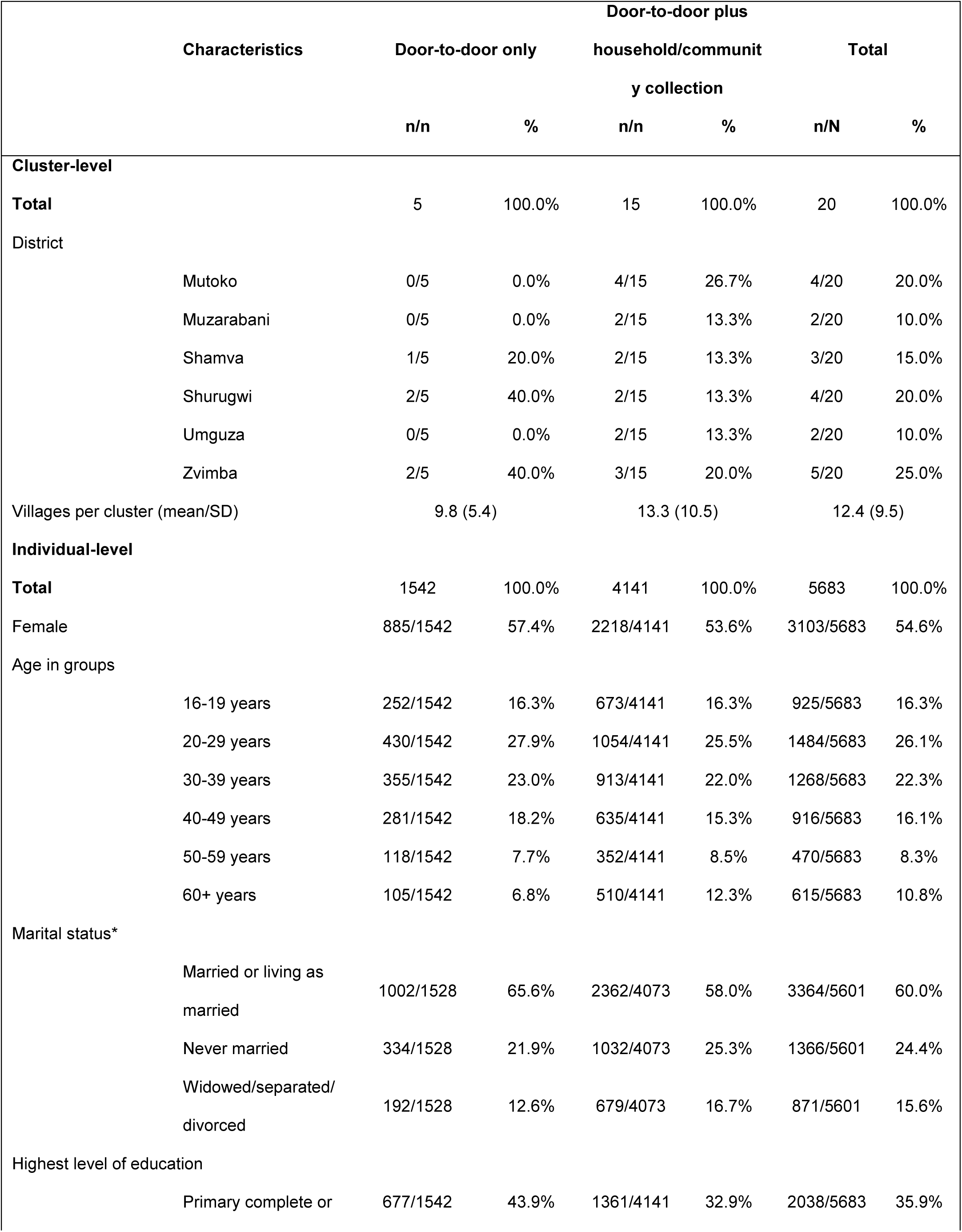
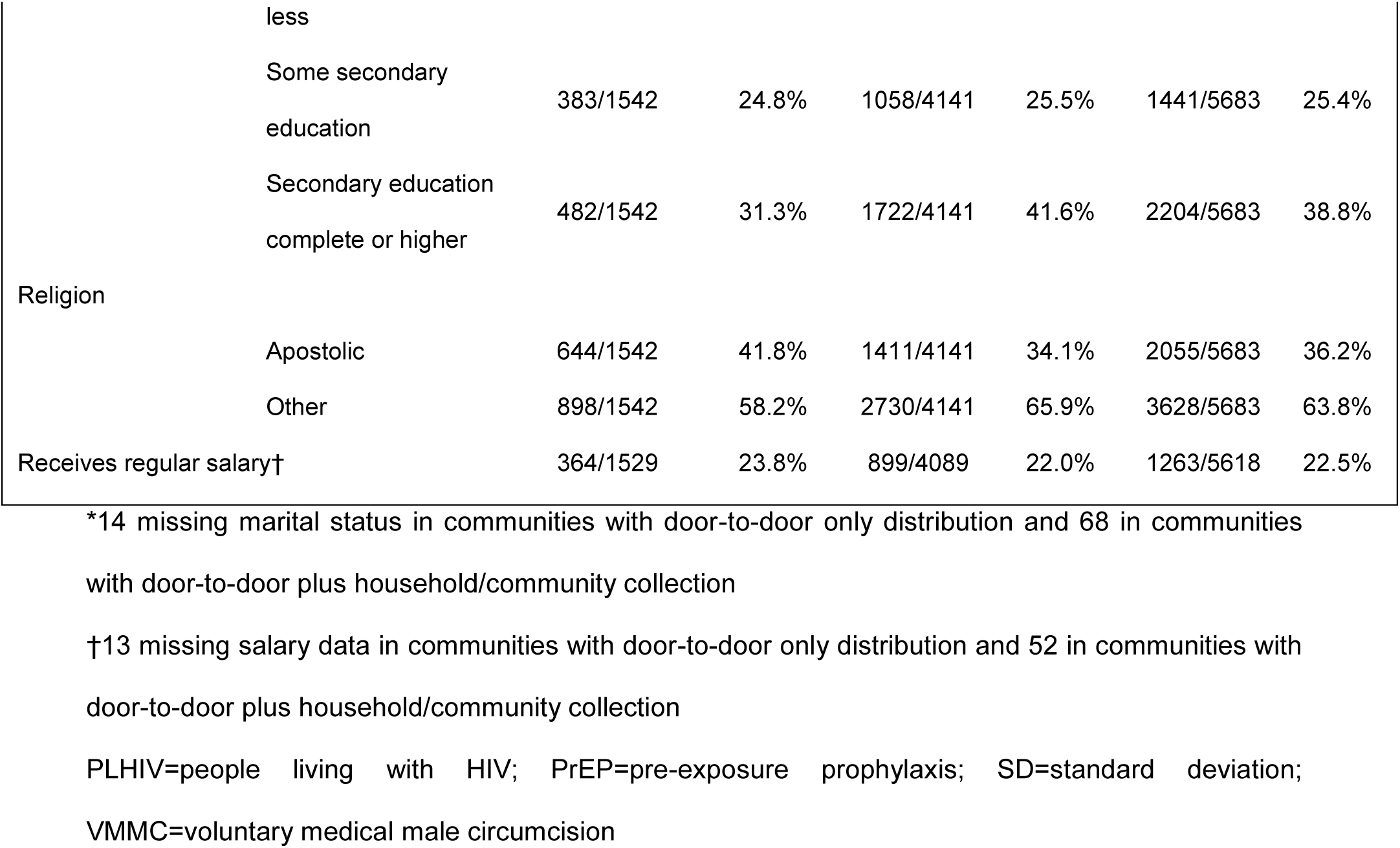
Cluster– and individual-level characteristics of participants in program and population-based survey by HIVST distribution model.

Based on participant observations, joint messaging on HIVST and U=U was well received during community engagement and was widely disseminated during distribution. Headman units actively participated in the design and implementation of their distribution models and were well-supported by local leaders before and during distribution.

From 3,000 households in headman units implementing community-led HIVST kit distribution, 5,683/6,748 eligible participants were surveyed, with a response rate of 84.2%. Among surveyed participants, 1,831 (32.2%) received a self-test kit of whom 1,229/1,831 (67.1%) reported using it, giving an overall HIVST uptake of 21.6% (1,229/5,683). Uptake did not differ by model of distribution: 358/1,542 (23.2%) in headman units implementing the door-to-door model compared to 871/4,141 (21.0%) for the combined model (adjusted odds ratio [aOR] 0.98 (95% confidence interval [CI]: 0.64-1.51, p=0.92).

### Participant and community characteristics

Tables 2-4 show cluster– and individual-level characteristics of participants in the programme and population-based survey by distribution model, levels of community involvement in planning distribution programmes, and community cohesion respectively. Participant characteristics were largely comparable by distribution model, levels of community involvement and cohesion group.

**Table 3.**
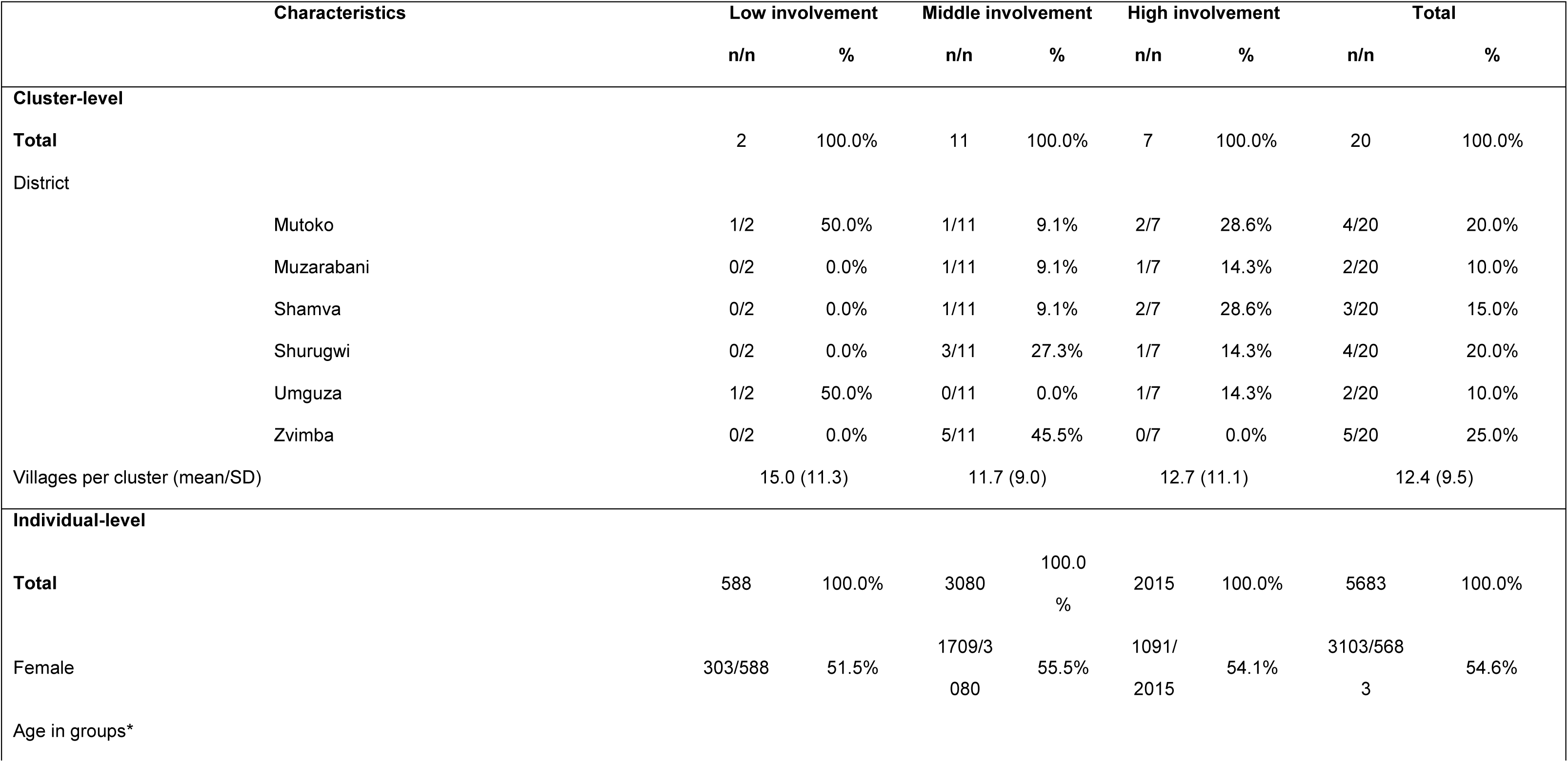
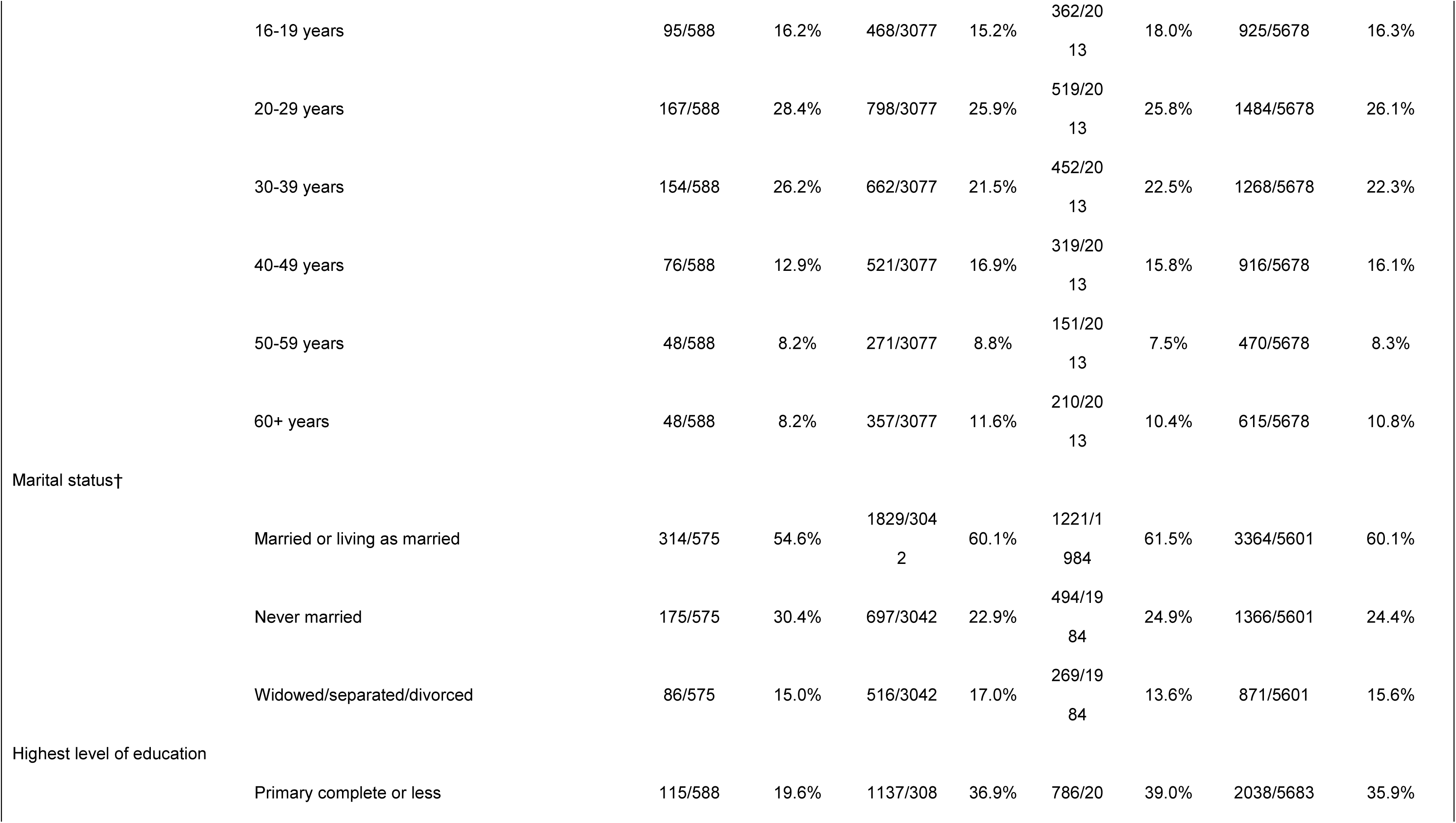
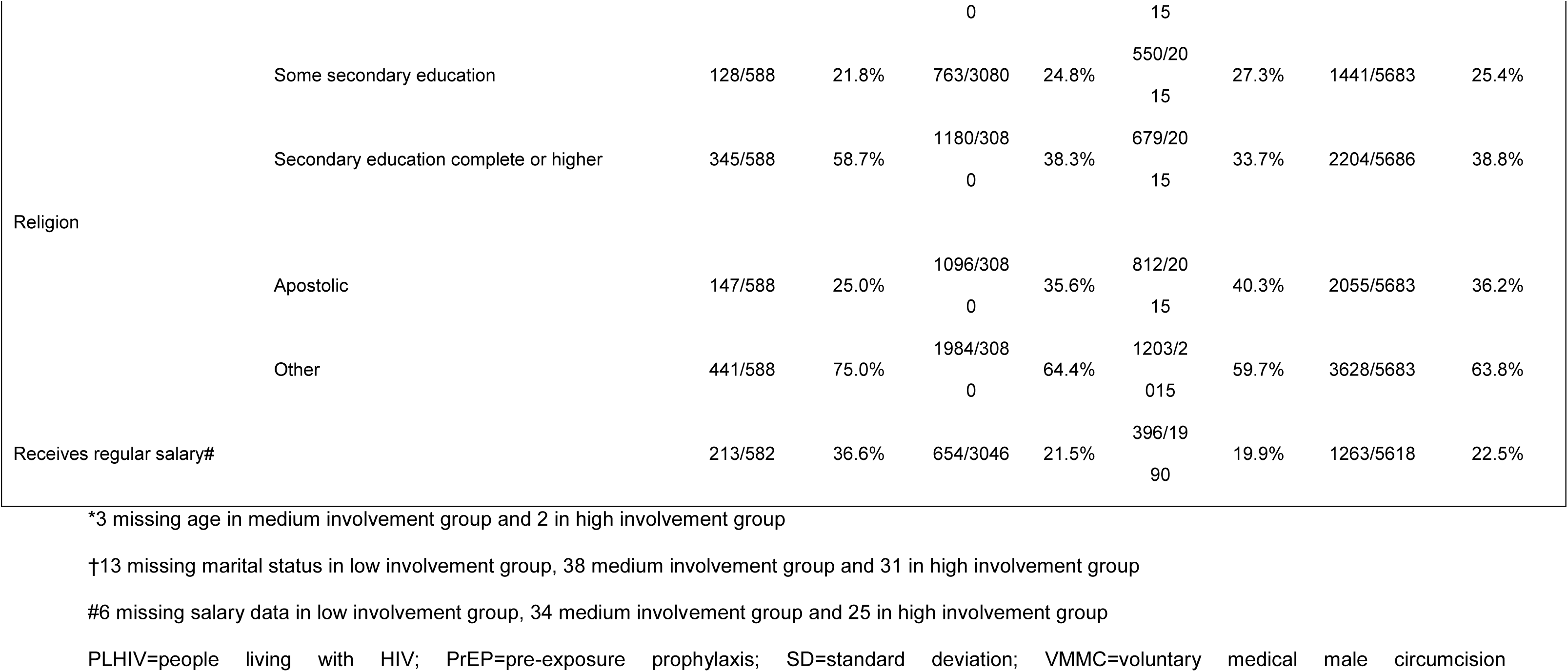
Cluster– and individual-level characteristics of participants in program and population-based survey by level of community involvement in planning distribution programs.

**Table 4.** Cluster– and individual-level characteristics of participants in program and population-based survey by self-reported community cohesion.

| Characteristics |  | Low cohesion |  | Middle cohesion |  | High cohesion |  | Total |  |
| --- | --- | --- | --- | --- | --- | --- | --- | --- | --- |
|  |  | n/n | % | n/n | % | n/n | % | n/n | % |
| Cluster-level |  |  |  |  |  |  |  |  |  |
| Total |  | 6 | 100.0% | 7 | 100.0% | 7 | 100.0% | 20 | 100.0% |
| District |  |  |  |  |  |  |  |  |  |
|  | Mutoko | 1/6 | 16.7% | 1/7 | 14.3% | 0/7 | 0.0% | 2/20 | 10.0% |
|  | Muzarabani | 0/6 | 0.0% | 2/7 | 28.6% | 1/7 | 14.3% | 3/20 | 15.0% |
|  | Shamva | 0/6 | 0.0% | 2/7 | 28.6% | 2/7 | 28.6% | 4/20 | 20.0% |
|  | Shurugwi | 4/6 | 66.7% | 1/7 | 14.3% | 0/7 | 0.0% | 5/20 | 25.0% |
|  | Umguza | 1/6 | 16.7% | 1/7 | 14.3% | 0/7 | 0.0% | 2/20 | 10.0% |
|  | Zvimba | 0/6 | 0.0% | 0/7 | 0.0% | 4/7 | 57.1% | 4/20 | 20.0% |
| Villages per cluster (mean/SD) |  | 8 (2.8) |  | 10.3 (11.5) |  | 18.3 (8.9) |  | 12.4 (9.5) |  |
| Individual-level |  |  |  |  |  |  |  |  |  |
| Total |  | 1770 | 100.0% | 1871 | 100.0% | 2042 | 100.0% | 5683 | 100.0% |
| Female |  | 992/1770 | 56.0% | 947/1871 | 50.6% | 1164/2042 | 57.0% | 3103/5683 | 54.6% |
| Age in groups |  |  |  |  |  |  |  |  |  |
|  | 16-19 years | 263/1770 | 14.9% | 326/1871 | 17.4% | 336/2042 | 16.5% | 925/5683 | 16.3% |
|  | 20-29 years | 475/1770 | 26.8% | 477/1871 | 25.5% | 532/2042 | 26.1% | 1484/5683 | 26.1% |
|  | 30-39 years | 439/1770 | 24.8% | 440/1871 | 23.5% | 389/2042 | 19.0% | 1268/5683 | 22.3% |
|  | 40-49 years | 293/1770 | 16.6% | 293/1871 | 15.7% | 330/2042 | 16.2% | 916/5683 | 16.1% |
|  | 50-59 years | 142/1770 | 8.0% | 144/1871 | 7.7% | 184/2042 | 9.2% | 470/5683 | 8.3% |
|  | 60+ years | 158/1770 | 8.9% | 189/1871 | 10.1% | 268/2042 | 13.1% | 615/5683 | 10.8% |
| Marital status* |  |  |  |  |  |  |  |  |  |
|  | Married or living as married | 1093/1748 | 62.5% | 1140/1840 | 62.0% | 1131/2013 | 56.2% | 3364/5601 | 60.1% |
|  | Never married | 409/1748 | 23.4% | 447/1840 | 24.3% | 510/2013 | 25.3% | 1366/5601 | 24.4% |
|  | Widowed/separated/divorced | 246/1748 | 14.1% | 253/1840 | 13.8% | 372/2013 | 18.5% | 871/5601 | 15.6% |
| Highest level of education |  |  |  |  |  |  |  |  |  |
|  | Primary complete or less | 612/1770 | 34.6% | 743/1871 | 39.7% | 683/2042 | 33.4% | 2038/5683 | 35.9% |
|  | Some secondary education | 405/1770 | 22.9% | 515/1871 | 27.5% | 521/2042 | 25.5% | 1441/5683 | 25.4% |
|  | Secondary education complete or higher | 753/1770 | 42.5% | 613/1871 | 32.8% | 838/2042 | 41.0% | 2204/5683 | 38.8% |
| Religion |  |  |  |  |  |  |  |  |  |
|  | Apostolic | 646/1770 | 36.5% | 746/1871 | 39.9% | 663/2042 | 32.5% | 2055/5683 | 36.2% |
|  | Other | 1124/1770 | 63.5% | 1125/1871 | 60.1% | 1379/2042 | 67.5% | 3628/5683 | 63.8% |
| Receives regular salary† |  | 409/1754 | 23.3% | 393/1846 | 21.3% | 461/2018 | 22.8% | 1263/5618 | 22.5% |

\*22 missing marital status in low cohesion group, 31 medium cohesion group and 29 in high cohesion group
†16 missing salary data in low cohesion group, 25 medium cohesion group and 24 in high cohesion group
PLHIV=people living with HIV; PrEP=pre-exposure prophylaxis; SD=standard deviation; VMMC=voluntary medical male circumcision

For community involvement in planning HIVST distribution programmes, 2 headman units were classified as low involvement, 11 were classified as medium involvement and 7 high involvement (Table 3). Headman units in the low involvement group were in 2 study districts, while those in medium and high involvement groups were spread across 5 study districts each.

For community cohesion, 6 headman units were in the low cohesion group and 7 headman units each in medium and high cohesion, respectively (Table 4). Headman units in low and high cohesion groups were in 3 study districts, while those in the medium cohesion group were in 5 study districts.

### Effect of community cohesion on outcomes

We found self-reported new HIV diagnosis increased with community cohesion, from 32/1,770 (1.8%) in the lowest cohesion group to 40/1,871 (2.1%) in the medium group, aOR 2.94 (1.41-6.12, p=0.004) and 66/2,042 (3.2%) in the highest cohesion group, aOR 7.20 (2.31-22.50, p=0.001) (Table 5).

**Table 5.**
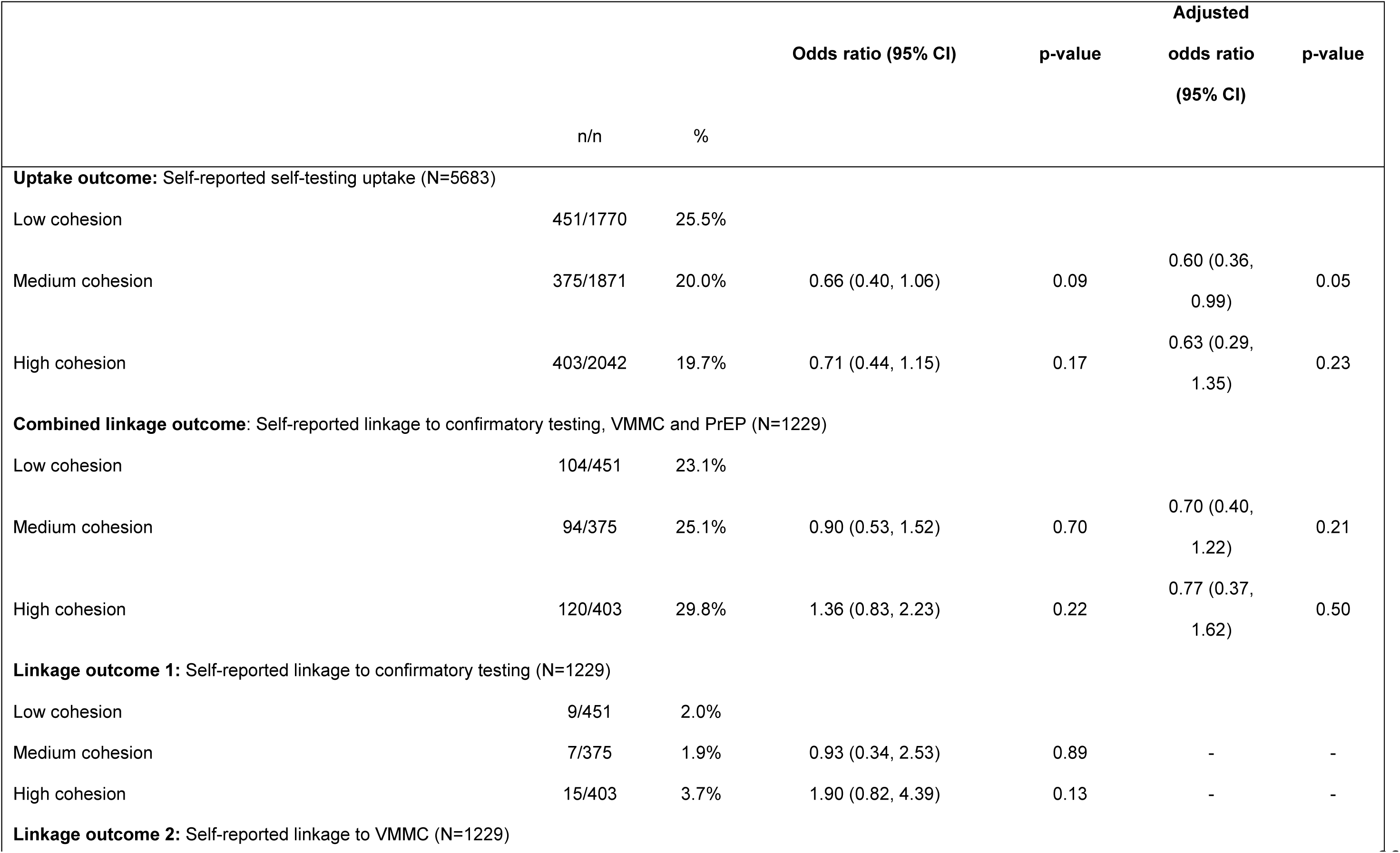
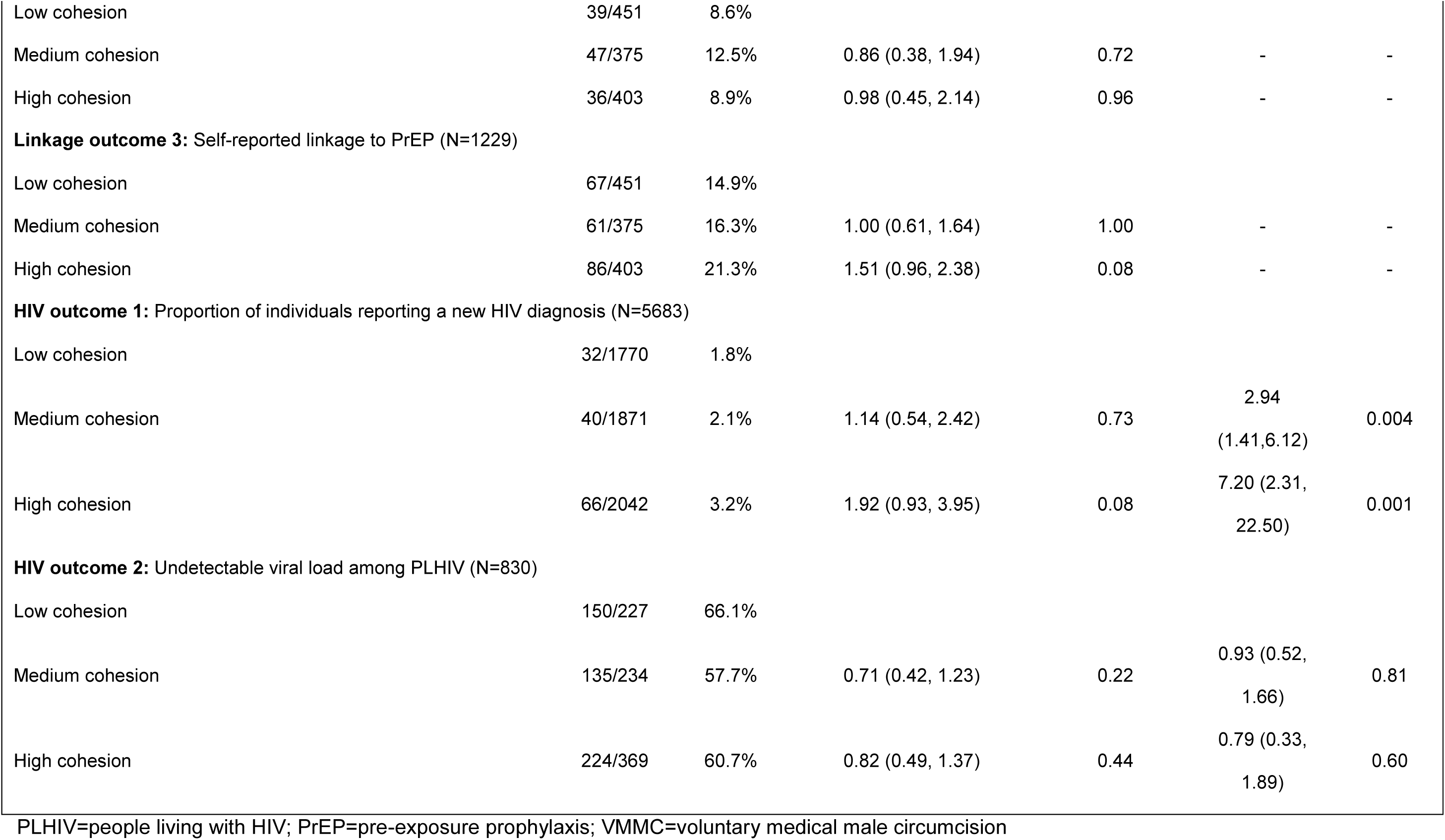
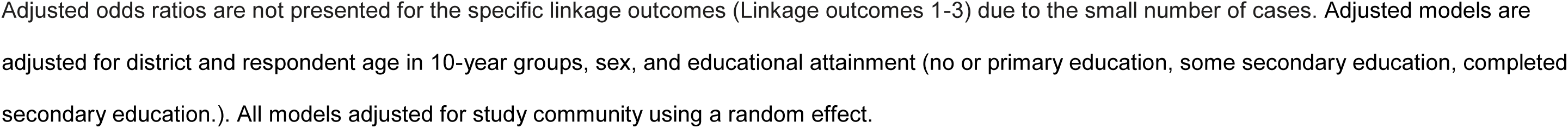
Comparison of outcomes by levels of community cohesion.

Other study outcomes did not differ by cohesion group. Cohesion had no overall effect on HIVST uptake across cohesion groups (p=0.42); 451/1,770 (25.5%) participants in the low group and 75/1,871 (20.0%) in the medium group (aOR 0.60 (0.36-0.99), p=0.05) reported uptake. In the high group, 403/2,042 (19.7%) participants (aOR 0.63 (0.29-1.35), p=0.23) reported HIVST uptake. Trend analysis using a linear parameterisation of the cohesion group variable showed there was no trend in cohesion and HIVST uptake; (aOR for 1-unit increase in cohesion score: 0.76 (95%CI: 0.51, 1.14), p=0.182). Similarly, there were no differences in linkage to post-test services across groups, with 104/451 (23.1%) participants linking in the low group, 94/375 (25.1%) in the medium group, (aOR 0.70 (0.40-1.22), p=0.21) and 120/403 (29.8%) in the high group (aOR 0.77 (0.37-1.62), p=0.50). Finally, undetectable viral load which was 150/227 (66.1%) participants in the low group, 135/234 (57.7%) in the medium group, (aOR 0.93 (0.52-1.66), p=0.81) and 224/369 (60.7%) in the high group (aOR 0.79 (0.33-1.89), p=0.60) did not differ.

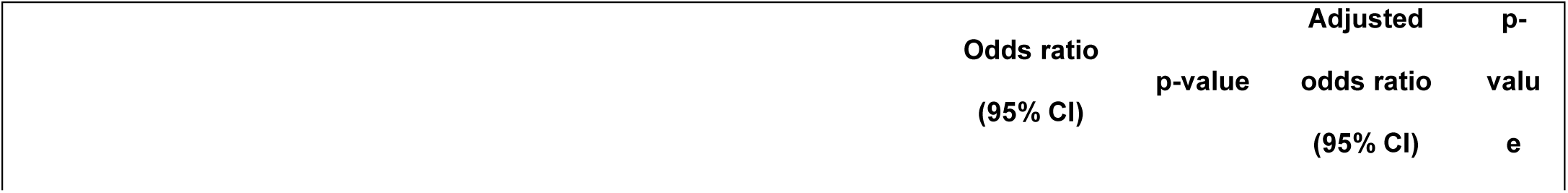

### Effect of HIVST distribution model on outcomes

Study outcomes did not differ by distribution model (Table 6).

**Table 6.**
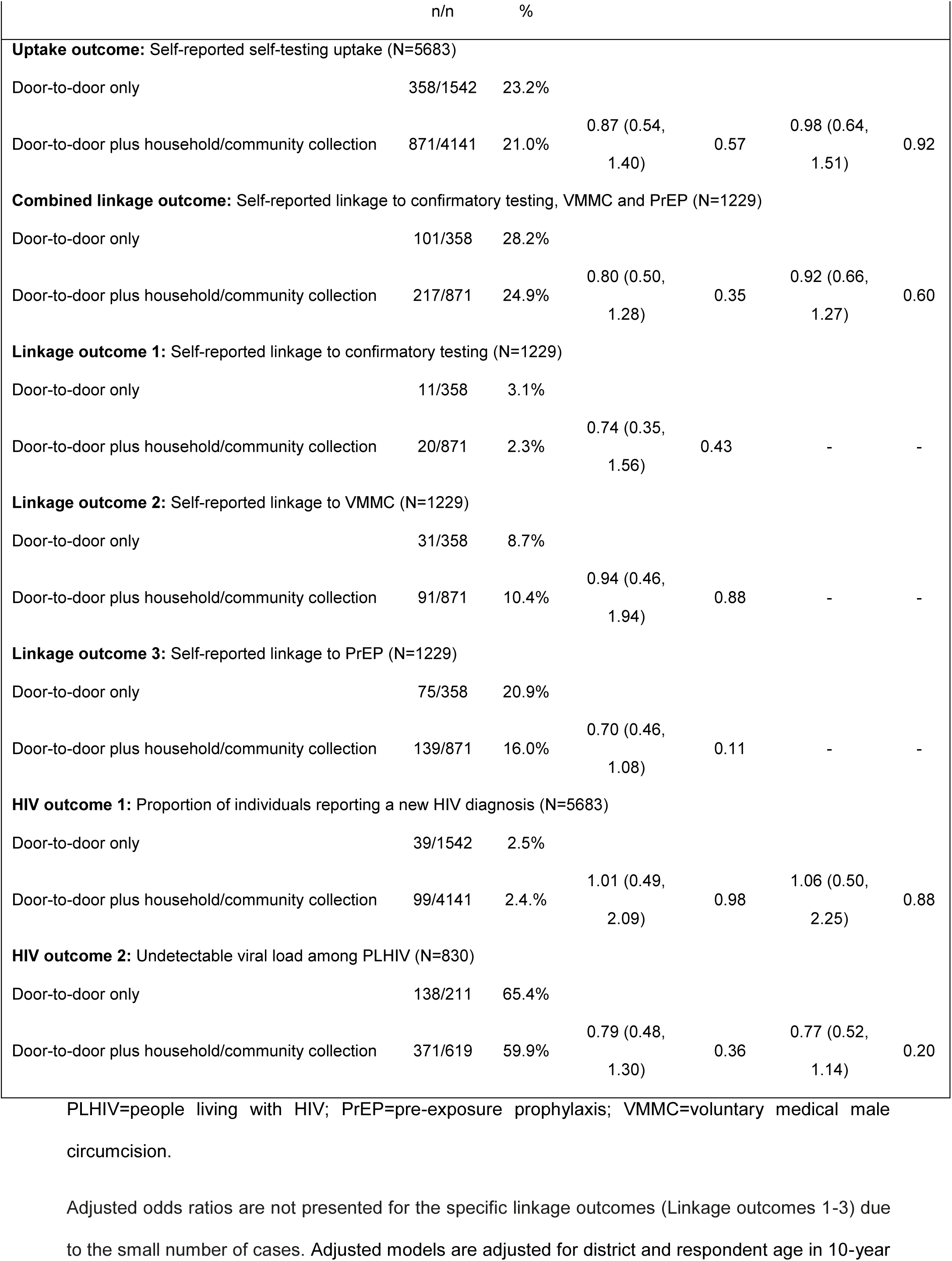
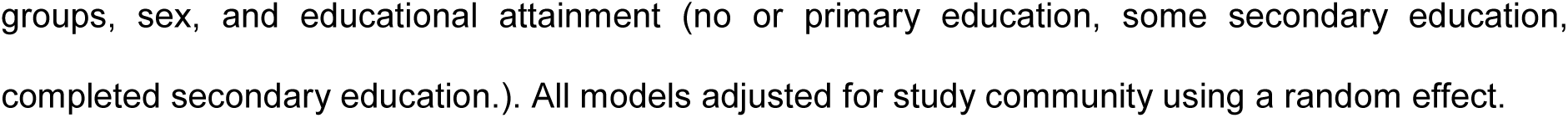
Comparison of outcomes by distribution model.

New HIV diagnosis was reported by 157/4,141 (3.8%) and 54/1,542 (3.5%) participants where combined or door-to-door distribution models were implemented, respectively (aOR 1.42 (95% CI: 0.79-2.54), p=0.24).

HIVST uptake in headman units implementing combined HIVST distribution models was reported by 871/4,141 (21.0%) participants and by 358/1,542 (23.2%) in headman units implementing door-to-door HIVST distribution only (aOR 0.98 (95% CI: 0.64-1.51), p=0.92). Independent of the distribution model and among all who received a self-test kit (1,831), use of the collected self-test kit did not differ by whether the kit was received door-to-door or elsewhere in the headman unit; 896/1,325 (67.6%) participants self-tested and received a kit door-to-door while 333/506 (65.8%) self-tested and received a kit by other means (aOR 1.08 (95% CI: 0.86-1.35), p=0.50). Similarly, at cluster-level self-testing uptake did not differ by whether the kit was received door-to-door or elsewhere in the headman unit; there was a – 2% change in HIVST uptake (95% CI –10, +7, p=0.72) in headman units implementing the combined HIVST distribution model compared with headman units conducting door-to-door distribution only. Linkage to post-test services was reported by 217/871 (24.9%) and 101/358 (28.2%) participants where combined or door-to-door distribution models were implemented, respectively (aOR 0.92 (95% CI: 0.66-1.27), p=0.60). Lastly, undetectable viral load was 371/619 (59.9%) and 138/211 (65.4%) among participants where combined or door-to-door distribution models were implemented, respectively (aOR 0.77 (95% CI: 0.52-1.14), p=0.20).

### Effect of levels of community involvement in planning distribution programmes on outcomes

Study outcomes did not differ by levels of community involvement in planning (Table 7).

**Table 7.**
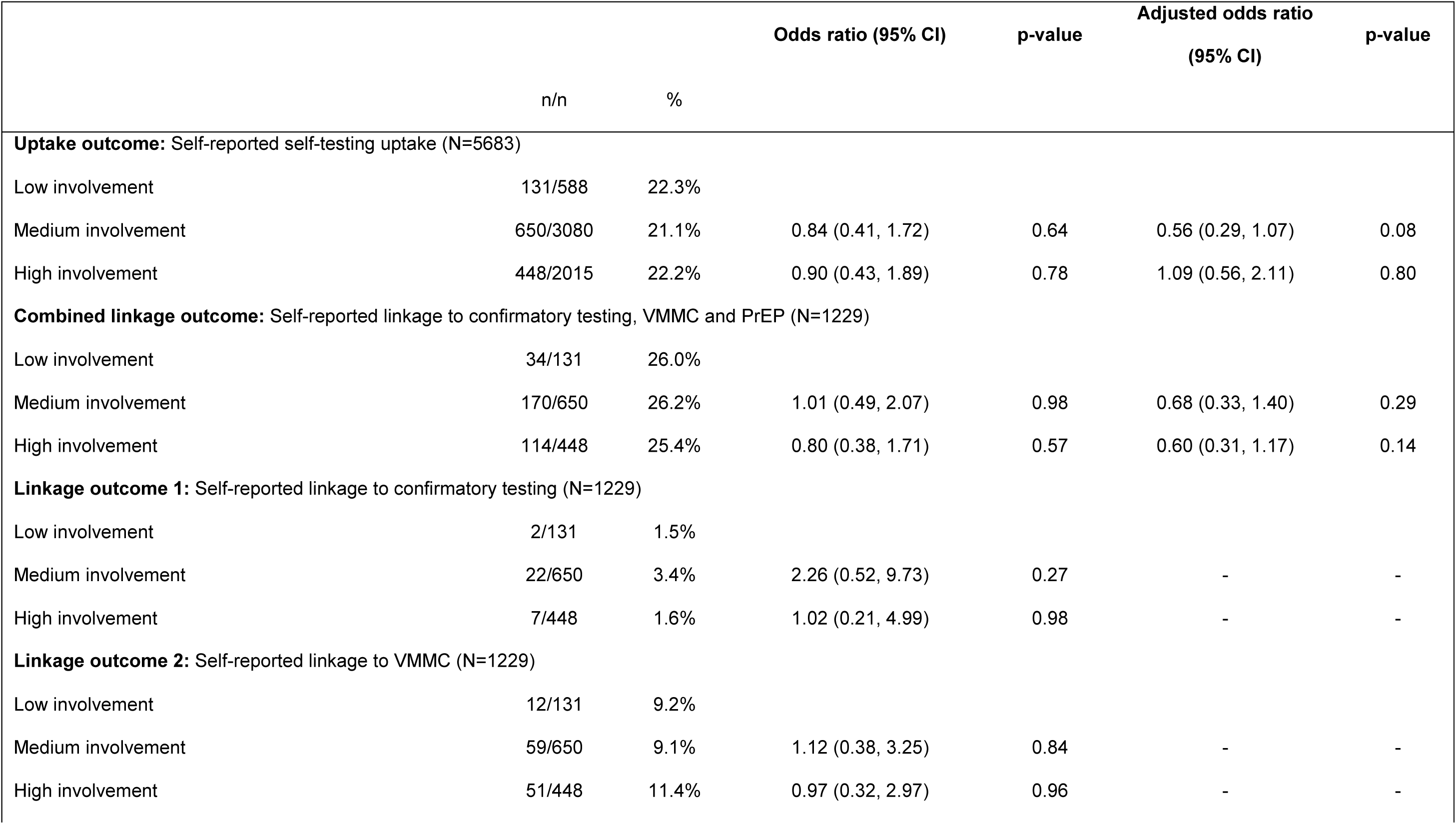
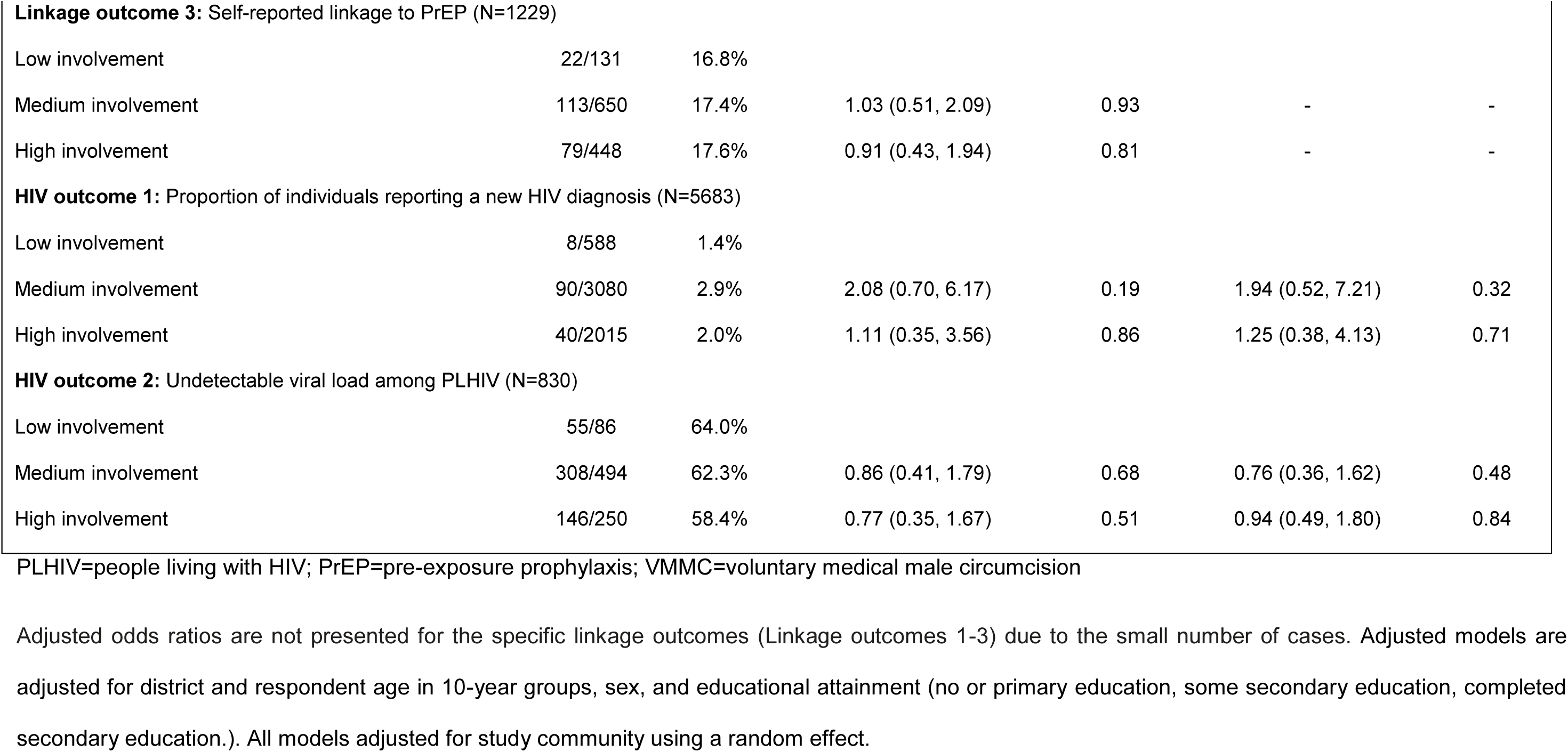
Comparison of outcomes by levels of community involvement in planning distribution programs.

|  | n/n | % | Odds ratio (95% CI) | p-value | Adjusted odds ratio<br>(95% CI) | p-value |
| --- | --- | --- | --- | --- | --- | --- |
| **Uptake outcome:** Self-reported self-testing uptake (N=5683) |  |  |  |  |  |  |
| Low involvement | 131/588 | 22.3% |  |  |  |  |
| Medium involvement | 650/3080 | 21.1% | 0.84 (0.41, 1.72) | 0.64 | 0.56 (0.29, 1.07) | 0.08 |
| High involvement | 448/2015 | 22.2% | 0.90 (0.43, 1.89) | 0.78 | 1.09 (0.56, 2.11) | 0.80 |
| **Combined linkage outcome:** Self-reported linkage to confirmatory testing, VMMC and PrEP (N=1229) |  |  |  |  |  |  |
| Low involvement | 34/131 | 26.0% |  |  |  |  |
| Medium involvement | 170/650 | 26.2% | 1.01 (0.49, 2.07) | 0.98 | 0.68 (0.33, 1.40) | 0.29 |
| High involvement | 114/448 | 25.4% | 0.80 (0.38, 1.71) | 0.57 | 0.60 (0.31, 1.17) | 0.14 |
| **Linkage outcome 1:** Self-reported linkage to confirmatory testing (N=1229) |  |  |  |  |  |  |
| Low involvement | 2/131 | 1.5% |  |  |  |  |
| Medium involvement | 22/650 | 3.4% | 2.26 (0.52, 9.73) | 0.27 | - | - |
| High involvement | 7/448 | 1.6% | 1.02 (0.21, 4.99) | 0.98 | - | - |
| **Linkage outcome 2:** Self-reported linkage to VMMC (N=1229) |  |  |  |  |  |  |
| Low involvement | 12/131 | 9.2% |  |  |  |  |
| Medium involvement | 59/650 | 9.1% | 1.12 (0.38, 3.25) | 0.84 | - | - |
| High involvement | 51/448 | 11.4% | 0.97 (0.32, 2.97) | 0.96 | - | - |
| **Linkage outcome 3: Self-reported linkage to PrEP (N=1229)** |  |  |  |  |  |  |
| Low involvement | 22/131 | 16.8% |  |  |  |  |
| Medium involvement | 113/650 | 17.4% | 1.03 (0.51, 2.09) | 0.93 | - | - |
| High involvement | 79/448 | 17.6% | 0.91 (0.43, 1.94) | 0.81 | - | - |
| **HIV outcome 1: Proportion of individuals reporting a new HIV diagnosis (N=5683)** |  |  |  |  |  |  |
| Low involvement | 8/588 | 1.4% |  |  |  |  |
| Medium involvement | 90/3080 | 2.9% | 2.08 (0.70, 6.17) | 0.19 | 1.94 (0.52, 7.21) | 0.32 |
| High involvement | 40/2015 | 2.0% | 1.11 (0.35, 3.56) | 0.86 | 1.25 (0.38, 4.13) | 0.71 |
| **HIV outcome 2: Undetectable viral load among PLHIV (N=830)** |  |  |  |  |  |  |
| Low involvement | 55/86 | 64.0% |  |  |  |  |
| Medium involvement | 308/494 | 62.3% | 0.86 (0.41, 1.79) | 0.68 | 0.76 (0.36, 1.62) | 0.48 |
| High involvement | 146/250 | 58.4% | 0.77 (0.35, 1.67) | 0.51 | 0.94 (0.49, 1.80) | 0.84 |

PLHIV=people living with HIV; PrEP=pre-exposure prophylaxis; VMMC=voluntary medical male circumcision
Adjusted odds ratios are not presented for the specific linkage outcomes (Linkage outcomes 1-3) due to the small number of cases. Adjusted models are adjusted for district and respondent age in 10-year groups, sex, and educational attainment (no or primary education, some secondary education, completed secondary education.). All models adjusted for study community using a random effect.

New HIV diagnosis did not differ across groups, with reports by 11/588 (1.9%) participants in the low group, 127/3,080 (4.1%) in the medium group, (aOR 1.98 (95% CI: 0.67-5.85), p=0.22) and 73/2,015 (3.6%) in the high group (aOR 1.73 (0.65-4.59), p=0.27).

There were no differences in HIVST uptake across community involvement groups, with 131/588 (22.3%) participants in the low involvement group, 650/3,080 (21.1%) in the medium involvement group, (aOR 0.56 (0.29-1.07), p=0.08) and 448/2,015 (22.2%) in the high involvement group (aOR 1.09 (0.56-2.11), p=0.80), reporting HIVST uptake. There were no differences in linkage to post-test services across groups, with 131/588 (22.3%) participants linking in the low group, 650/3,080 (21.1%) in the medium group, (aOR 0.56 (0.29-1.07), p=0.08) and 448/2,015 (22.2%) in the high group (aOR 1.09 (0.56-2.11), p=0.80). Undetectable viral load which was 55/86 (64.0%) among participants in the low group, 308/494 (62.3%) in the medium group, (aOR 0.76 (0.36-1.62), p=0.48) and 146/250 (58.4%) in the high group (aOR 0.94 (0.49-1.80), p=0.84) did not differ. Finally, there were no statistically significant associations between (i) distribution model and levels of community involvement (p=1.0), (ii) distribution model and community cohesion (p=0.13), and (iii) levels of community involvement and community cohesion (p=0.15).

## DISCUSSION

In this mixed-methods study, we examined the effects of community cohesion, HIVST distribution models and levels of community involvement in planning distribution programmes on: (i) self-reported new HIV diagnosis (ii) self-reported HIVST uptake; (iii) self-reported linkage to confirmatory testing, VMMC and PrEP; and (iv) viral load, among headman units conducting community-led HIVST kit distribution. We found the proportion of participants reporting a new HIV diagnosis increased with evidence of community cohesion and there was a dose response, with 1.8%, 2.1% and 3.2% in low, medium, and high cohesion groups respectively. The type of HIVST distribution models implemented by headman units did not affect outcomes, nor did levels of community involvement in planning.

The finding of self-reported new HIV diagnosis increasing with community cohesion is in line with our hypothesis that cohesive communities would achieve better outcomes. Similar evidence was found in the parent trial; in high cohesion communities the odds of new HIV diagnosis was greater in the community-led arm than in the comparison arm (OR 2.06 (95% CI: 1.03-4.19), p= 0.04) [21].

There is some evidence that cohesive communities are close-knit with a sense of social identity, belonging and understanding of each other‘s health needs. In addition, working together provides space to confront myths, misconceptions and stereotypes about people living with HIV (PLHIV) thereby reducing HIV stigma. In Zimbabwe, participation in community groups facilitated linkage to HIV prevention, care and treatment services and was associated with lower levels of HIV stigma [22,23] (the adverse effects of stigma on uptake of HIV-related services, health outcomes and quality of life among PLHIV has been documented [24,25,26]). In cohesive communities, community members’ concern for good health extends beyond the individual to other members. Guided by group-based norms and values – the belief that ―together we achieve better and more‖ (collective efficacy) – cohesive communities would collaborate effectively to achieve a common goal, eliminating new HIV infections through community-led HIVST distribution. In such headman units, U=U campaigns could have appealed to community members and motivated testing. As a result, HIVST kit distributors knew who to target with HIVST (active case finding), furthermore, good existing social relationships and trust for the distributor mediated community members’ acceptance of the offer of kits [15], resulting in people who would not otherwise test, opting to test.

The lack of differences in self-testing uptake by community cohesion and distribution model could be attributed to each headman unit working together to design and refine ways of distributing kits in their setting. Such models would overcome context-specific barriers to achieve optimal uptake.

Even though cohesion was associated with higher reports of new HIV diagnosis in this study and the HIVST distribution period was associated with higher ART initiation rates at health facilities with or without HIVST in their catchment areas [18], it is likely that those newly diagnosed and initiated on ART under WHO‘s ―Treat All‖ policy [27] may not have achieved undetectable viral load by the time of the survey (3-4 months after distribution). In their study Ali et al. found the median time to achieve viral load suppression after initiation of ART to be 181 days (CI: 140.5-221.4) [28]. This may explain why the other community measures (distribution models and levels of community involvement in planning) had no effect on undetectable viral load.

In the trial within which this study was nested, we found similar outcomes between the community-led and the paid distributor arms; linkage outcomes and reports of new HIV diagnosis in the intervention arm were comparable with those using a paid distributor model [18], showing communities were able to develop models that worked for them and optimised outcomes. Our process evaluation data (not presented here) suggests barriers to linkage to post-test services some of which have been reported in other studies; the belief that linkage to post-test services is unnecessary for HIV negative people [29,30,31], poor or inaccurate knowledge of PrEP [31,32,33,34] and VMMC [35,36], fear of pain during the VMMC procedure [35,36,37] and long distances to health facilities [38]. Finally, healthcare workers shared the view that linkage works better if they are incentivised for each client linking to services [39]; incentives were not provided in this study. These barriers may have affected linkage to post-test services in headman units for each of the community factors.

The strengths of this study included the use of robust methods for documenting and analysing how the community-led intervention was implemented in each community. This study adds to evidence on the positive effects of community cohesion on positive health behaviours and outcomes [15,17]. While most studies on social cohesion consider cohesion at the individual level [15,17], this study attempted to measure community cohesion systematically by observing levels of community involvement in planning HIVST distribution programmes using a structured observation tool (i.e., community-level). Our measure, levels of community involvement in planning, relates to 4 out of 6 characteristics of Campbell et al, 2013 [22] conceptualisation of HIV competent communities, namely: (i) critical thinking about obstacles to health-enhancing behaviour change, and discussions of locally realistic strategies for tackling these; (ii) promoting a sense of local ownership and responsibility for contributing to efforts that combat HIV/AIDS (iii) fostering a sense of solidarity and common purpose in confronting HIV/AIDS and (iv) identification of individual and group strengths for this challenge [22]. Furthermore, use of this measure was moderated by independent scoring by two researchers and resolving discrepancies through consensus.

Limitations of the study include the reliance on self-reports for study outcomes. While it is possible willingness to self-report could have varied by community cohesion, this is unlikely as this factor seemed to affect new HIV diagnosis but not other self-reported outcomes such as HIVST uptake or linkage. We tried to minimise self-reporting bias by using ACASI as well as laboratory testing of DBS samples. Levels of community involvement and community cohesion are related constructs, and the former may be a feature of community cohesion. However, the community involvement variable was weakly associated with the validated community cohesion measure – possibly due to the small sample size of 20 communities – and the associations between community cohesion and new HIV diagnosis were in line with our hypothesis at the beginning of the study. Although we systematically measured community involvement using participant observation, this was not a validated approach.

In summary, in this mixed-methods study we found community-led interventions to distribute HIVST kits are feasible and acceptable among rural Zimbabwean communities, accommodating flexibility in design of community-led HIVST kit distribution models and varying levels of community involvement in planning distribution to achieve outcomes similar to those in programmes that were implemented by professionally supported, paid distributors. Community cohesion in rural settings was associated with an increase in self-reported new HIV diagnoses. This suggests that more cohesive communities may be better able to identify those most at risk of undiagnosed HIV infection, and that in closely-knit communities people who need to test are more likely to accept the offer of self-test kits from fellow community members, under a programme that is validated by their community leaders and in an environment where HIV issues can be discussed freely. Qualitative research may provide further insights and be used to improve community-led HIVST programmes as distribution is scaled-up. Regardless of levels of community cohesion, future community-led HIVST programmes may be implemented successfully by enhancing messaging on HIVST and post-test services; addressing related knowledge gaps; and confronting HIV-related myths, misconceptions, and stereotypes (stigma reduction interventions). Communities can learn from and adopt these participatory community-led approaches to intervention planning and implementation for HIVST and other health priorities they identify. Continued implementation of community-led interventions may increase community cohesion and benefit various public health programmes.

## Author contributions

Conceptualization: MN, MT, CCJ, KH, ELC, FMC, ELS. Data curation: MKT, NR, MN, CW, MM, MT, ELS. Formal analysis: MKT, NR, MN, CW, MT, ELS. Funding acquisition: MT, CCJ, KH, ELC, FMC, ELS. Investigation: MKT, NR, CW, MM. Methodology: MKT, NR, CW, MM, MT, ELS, FMC, ELS. Project administration: MKT, NR, CW, MM, ELS. Writing – original draft preparation: MKT, NR, MN. Writing – review and editing: MKT, NR, MN, MT, CCJ, FMC, ELS.

## Data Availability

Data are available upon request. De-identified data are available from the corresponding author upon request.

## Acknowledgements

The authors appreciate the support of the Zimbabwe Ministry of Health and Child Care and the STAR Initiative Consortium partners, and the contribution of field researchers and participants, who all made this study possible.

